# Long-term outcome of corneal collagen crosslinking with riboflavin and UV-A irradiation for keratoconus

**DOI:** 10.1101/2022.02.05.22270507

**Authors:** Franziska K. Seifert, Johanna Theuersbacher, Dorothee Schwabe, Olga S. Lamm, Jost Hillenkamp, Daniel Kampik

## Abstract

**Purpose:** To evaluate long-term outcomes of corneal collagen crosslinking (CXL) using riboflavin and UV-A irradiation and to determine when to repeat CXL.

**Methods:** In this retrospective consecutive interventional case series 131 eyes of 131 patients (95 male, 36 female, mean age 29.7±11.4 years) between 2006 and 2016 received standard CXL (Dresden protocol, epithelium-off) for progressive keratoconus. Corrected distance visual acuity (CDVA) and corneal tomography (K_1_, K_2_, K_max_) were repeatedly recorded 1 year (n=103 eyes) to 10 years (n=44) postoperatively. Only one eye per patient was included. Paired t-test or Wilcoxon matched-pairs signed rank test was used for parametric and nonparametric data, respectively.

**Results:** 1 to 3 years *pre*operatively, median K_2_ significantly increased by 1.1D (p<0.001). *Post*operatively, median K_2_ increased by 0.1D after 1 year, then decreased over the remaining postoperative period by 0.85D (p=0.021). Median apical corneal thickness decreased by 16µm (p=0.012) after 5 years and then returned to preoperative values. Mean CDVA showed a significant improvement (decrease in logMAR 0.12 after 10 years, p=0.010). K_max_ fluctuated without significant change. CXL non-responders, defined by a postoperative increase in K_max_>2D, increased from 16% after 5 to 33% after 10 years. Risk factors for non-response were young age, high astigmatism, thin cornea, and atopic dermatitis. 4 eyes were re-treated 3–4 years after first CXL without complications and keratoconus stabilized thereafter.

**Conclusions:** CXL can slow or stop keratoconus progression. However, as the number of responders declines after 5 years, patients with risk factors may require re-treatment.

## Introduction

Keratoconus is a non-inflammatory bilateral, progressive ectasia in which the cornea is thinned, loses stability, and thus becomes progressively conical in shape^1^. Lamellar or penetrating keratoplasty is the only option for restoring visual acuity in advanced stages^2^. In the past 15 years, a less invasive therapy with corneal collagen crosslinking (CXL) has become available. The aim of this treatment is to induce crosslinks between collagen lamellae of the corneal stroma using UV-A light and riboflavin as a photosensitizer. This leads to an increase in corneal stability and thus prevents or slows down further progression^3-6^. In case of recurrent disease progression, this procedure can be repeated. In 2003, Wollensak et al. published the first results of CXL treatment for keratoconus^7,8^. In the meantime, several studies have described the efficacy and safety of the intervention in the medium term (3 - 7 years)^9-12^. Although CXL has become a standard procedure for treating progressive keratoconus, only a few long-term studies have been conducted. To date, Raiskup et al., Theuring et al., and Nicula et al. have published results with a 10-year follow-up^12-14^. Recently, Vinciguerra et al. published a study with a mean follow-up period of 11 years and a maximum of 13 years^15^. All of these studies showed a decrease in corneal curvature and astigmatism and an improvement in visual acuity.

Aim of this study was to assess the long-term safety and efficacy of standard CXL (Dresden protocol) for progressive keratoconus and to identify risk factors for recurrence after an initial successful CXL.

## Materials and Methods

### Study design

This retrospective study included patients with progressive keratoconus treated with CXL at our hospital between 2006 and 2016. Data were collected from patients with a follow-up period of at least 1 year and a maximum of 10 years. Data collection was approved by the University’s ethics committee (ref. no. 89/08) and meets the principles of the Declaration of Helsinki. A total of 131 eyes of 131 patients were included in the study. Indication criteria for CXL were an increase in K_max_ (steepest curvature value based on the sagittal (axial) anterior curvature map) by over 1 D within 6 months, an increase in astigmatism or myopia by at least 1 D within 1 year, and a corneal thickness of more than 400 µm before abrasion. CXL was not performed in advanced keratoconus with a K_max_ value > 70 D.

Data were collected consecutively, resulting in a decreasing number of patients over time. In addition, some patients showed a patchy follow-up. In 44 patients, a follow-up period of 10 years was achieved.

### Assessments

To assess disease progression before surgery, data were collected 1-3 years and 3-9 months preoperatively. The Efficacy of CXL in terms of changes in spectacle-corrected distance visual acuity (CDVA) in logMAR, corneal curvature (K_max_ and K_2_), and apical corneal thickness measured using Scheimpflug tomography (Pentacam HR, Oculus, Germany) was determined 6 months, 1 year, and 2, 3, 5, 7, and 10 years postoperatively. Slitlamp findings were assessed for typical changes, such as Vogt striae, corneal scars, and iron lines. A contact lens-free period of at least 48 h before examination was maintained. In addition, the safety of the treatment was assessed based on the occurrence of complications during long-term follow-up.

### Treatment

CXL was performed according to the Dresden standard protocol. The epithelium, with a diameter of 8 mm, was removed under topical anaesthesia (oxybuprocain) using a blunt hockey knife. Subsequently, riboflavin eye drops (isotonic 0.1% riboflavin solution with 20% dextran) were applied to the cornea every 3 minutes for 30 minutes. Before UV-A irradiation, the diffusion of riboflavin into the anterior chamber was detected using a slit lamp with a blue filter. Irradiation was carried out for 30 min at a wavelength of 370 nm, intensity of 3 mW/cm^2^, and diameter of 8 mm, with further application of riboflavin every 5 min (UVX 1000, IROC Innocross AG, Zug, Switzerland). After treatment, patients received a soft bandage contact lens and antibiotic eye drops (ofloxacin) three times daily for one week and steroid eye drops (dexamethasone) twice daily for two weeks.

### Statistical analysis

SPSS was used for statistical data analysis (IBM, Version 24.0 and 25.0). Pre- and postoperative changes were measured using the paired *t*-test or Wilcoxon matched-pairs signed-rank test for parametric and nonparametric data, respectively. Normal distribution was tested using the Shapiro-Wilk test. Because the right and left eyes in keratoconus represent dependent samples, only one eye per patient was included for correct statistical analysis. If CXL was also performed on the fellow eye, only the first eye with CXL was included.

For comparisons between groups, the *t*-test for independent samples or Mann-Whitney *U* test was used for parametric and nonparametric data, respectively. If the data to be compared were categorical, both groups were compared using cross tables and the chi-squared test or Fisher’s exact test. To determine the risk factors for therapy failure, patients were divided into two groups, responders and non-responders, depending on postoperative changes in K_max_. Risk factors were evaluated using binary logistic regression analysis and odds ratios.

## Results

Between 2006 and 2016, 167 patients underwent CXL for keratoconus at our hospital. 36 patients were excluded from the study: in 26 patients, the follow-up time was <1 year; in one patient, topographic follow-up was not possible due to a lack of compliance; in one patient, irradiation time deviated from the standard protocol and in five other patients, the corneal thickness was < 400 µm preoperatively; two patients had high preoperative K_max_>70 D; one patient did not agree with data collection. A total of 131 patients (male:female ratio = 95:36) were included in the study. Mean age was 29.7 (± 11.4) years, male patients were on average younger (27.2 ± 9.8 years) than female patients (36.3 ± 12.7 years). At the time of the procedure, the oldest patient was 63 years old, and the youngest patient was 11 years old.

Due to the young patient cohort and the long observation period of 10 years, some time points were missed. To prevent this from distorting the data series, we opted for a pairwise comparison of pre- and postoperative parameters at each time point, allowing the use of paired statistical tests. This allows the comparison of different patient numbers at different time points over 10 years but also prevents the addition of error bars to each time point in a time course diagram (Fig. 1). To plot the variation and error bars, see Supplementary Fig. 1 which shows box plots comparing pre- and postoperative parameters separately for each time point.

**Fig. 1.**
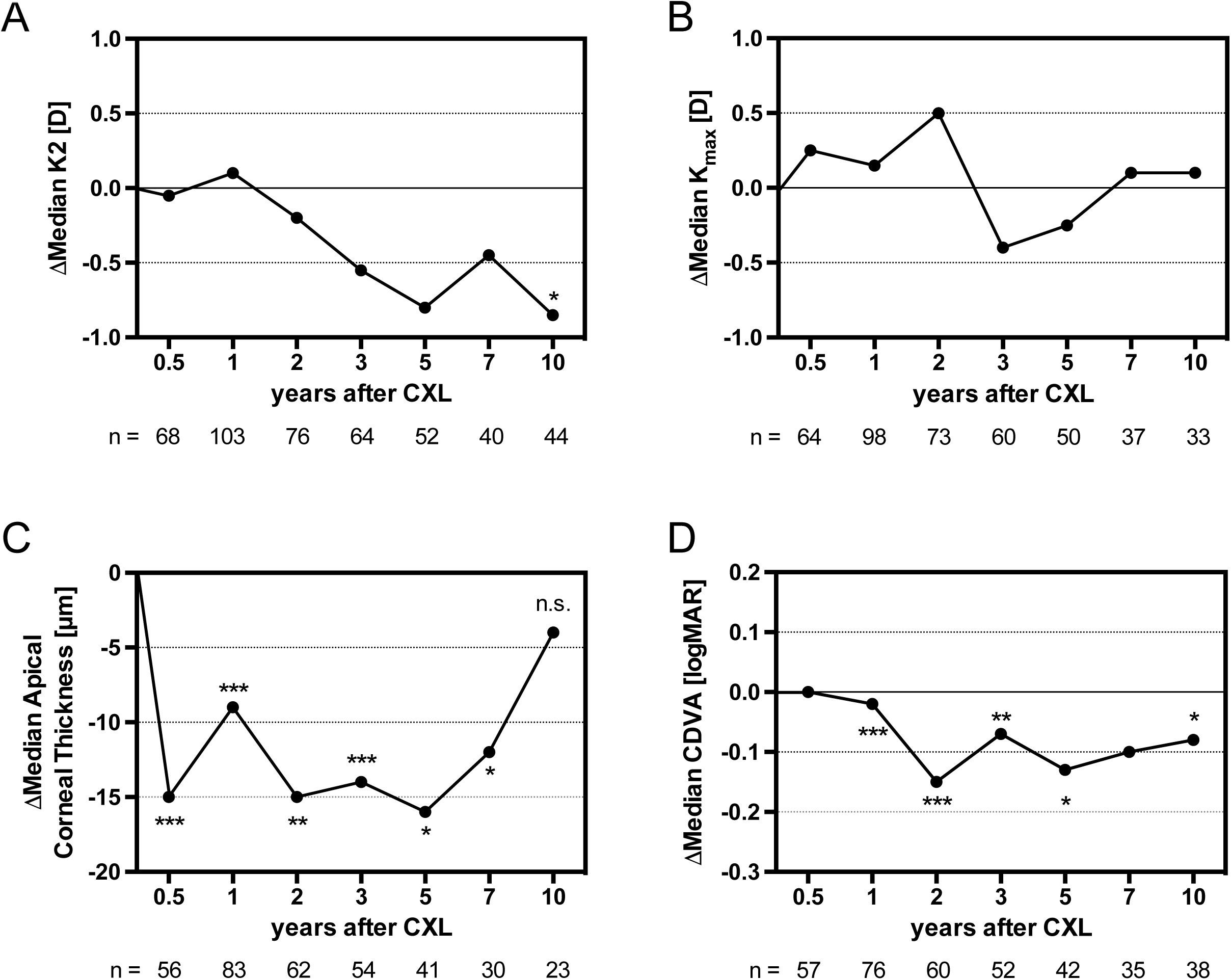
Development of parameters indicating keratoconus progression after corneal collagen cross-linking (CXL). Because differences of medians are calculated from pairs (postop minus preop), it is not legitimate to plot standard deviation or other indicators of variation. (a) Median K_2_ change: Up to 1 year postoperatively, a slight increase of curvature was observed. Beyond 1 year, K_2_ decreased by up to 0.85 D at 10 years (*: p < 0.05, all other data points: not significant). (b) Median K_max_ change: Fluctuations without significant changes were observed over the entire follow-up period. (c) Median corneal thickness at the apex: Beyond 6 months postoperatively, a significant stabilization without any further thinning tendency was observed (***: p<0,001, **p < 0.005, *: p < 0.05, n.s., not significant). (d) Change of median corrected distance visual acuity (CDVA): Beyond 2 years after treatment, an improvement of CDVA by about one line was observed (***: p<0,001, **p < 0.005, *: p < 0.05).

During the *preoperative* period, keratoconus progression was observed in all eyes. Median K_2_ and K_max_ both showed a significant increase of 1.1 D (p<0.001) and 1.5 D (p=0.001), respectively, during preoperative years 2 to 3. Nine to three months preoperatively, both K_2_ and K_max_ increased by 1.4 D (p<0.001) and 1.0 D (p=0.007) respectively. Apical corneal thickness decreased on average by 12.0 ±28.9 µm (p=0.003) during preoperative year 2 to 3, and by 4 ±31,3 µm (p=0.064) at nine to three months preoperatively. Best-corrected visual acuity did not show any significant changes.

*Postoperatively*, median K_2_ did not change in the first year, but a continuous decrease by 0.8 D (p=0,485) after 5 years was observed. 10 years after CXL, median K_2_ has decreased by 0.85 D (p=0.021) in comparison to the preoperative baseline (Fig. 1a). Median K_max_ showed high fluctuations over the entire follow-up period without any significant changes or trends (Fig. 1b). After 2 years, it increased by 0.5 D (p=0.311), and after 7 and 10 years the median increased by approximately 0.1 D (p=0.526 and p=0.276). Apical corneal thickness decreased postoperatively. After 2 and 5 years, the median values were 15 µm (p=0.002) and 16µm (p=0.012) below the initial value, respectively. From 5 years onwards, a tendency toward thickening was observed. 10 years after CXL, apical corneal thickness was not statistically different from the preoperative baseline (p=0.487) (Fig. 1c). CDVA in logMAR improved in the postoperative course at all follow-up time points by approximately one line. 10 years after CXL, a significant decrease in CDVA of 0.08 (p=0.010) was observed (Fig. 1d). The proportion of patients with an improvement of at least one line was always higher than that of patients with a loss of at least one line. After 2 and 3 years, 53 % and 48 % showed an improvement in visual acuity, whereas 13 % and 15 % showed a deterioration. After 7 and 10 years, 80 % and 84 % showed improvement or stabilization of CDVA (Fig. 2).

**Fig. 2:**
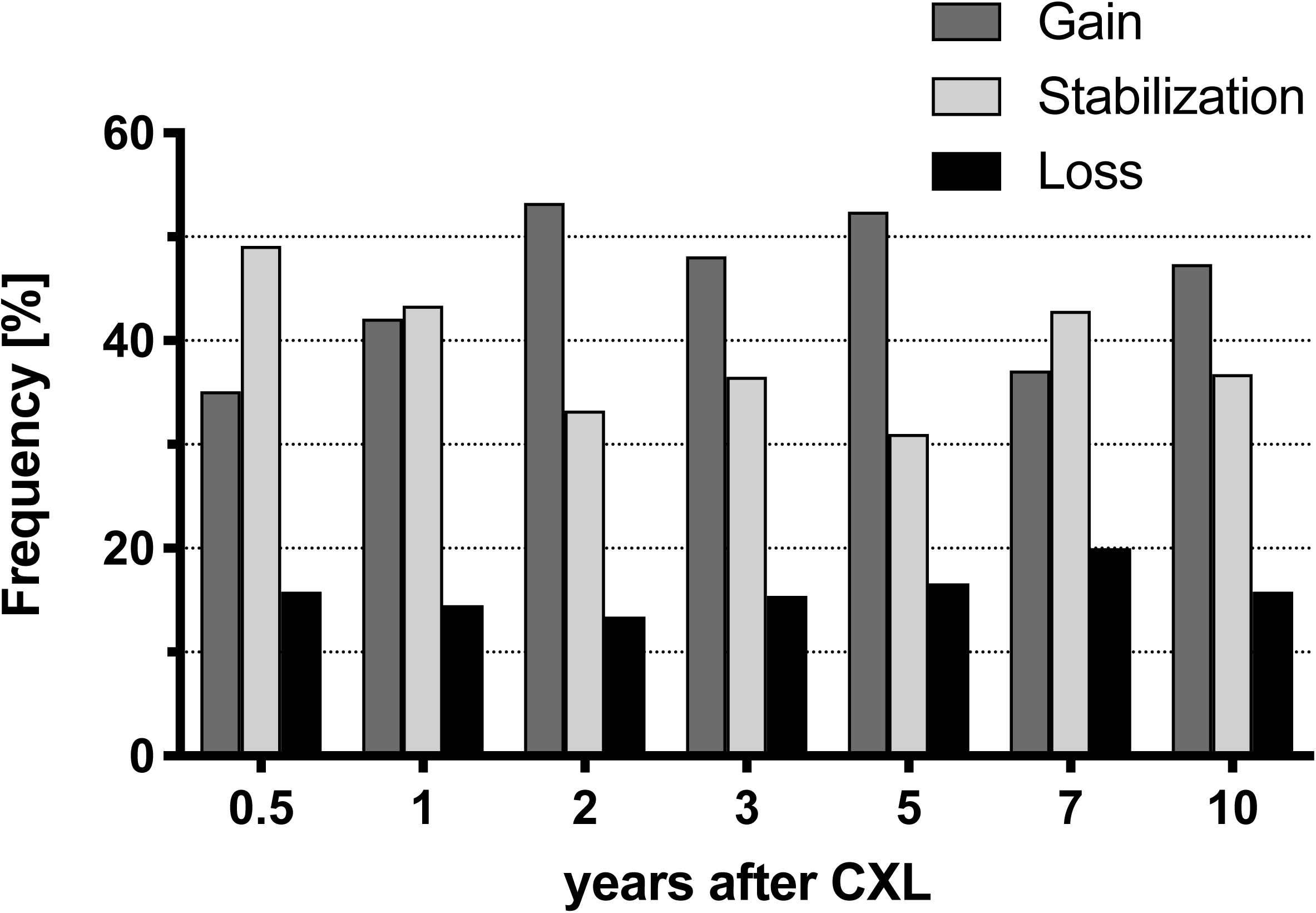
Postoperative changes in corrected distance visual acuity (loss of CDVA: increase by at least 0.1 logMAR levels; improvement of CDVA: decrease by at least 0.1 logMAR levels). The number of patients with stable vision or improvement of at least one line (grey bars) was significantly higher over the entire follow-up period than the number of patients who showed a loss of at least one line (solid black bars).

To determine which patients benefited from CXL therapy and who did not, they were divided into two groups: non-responders were identified by a postoperative increase of K_max_ ≥ 2 D, all others with a postoperative stabilisation of K_max_ were defined as responders, that is, an increase of < 2 D or any decrease. Beyond 5 years after treatment, the non-responder rate increased from 16 % after 5 years to 33 % after 10 years (Fig. 3). To identify the risk factors for disease recurrence, preoperative factors were compared between the two groups: high astigmatism, thin cornea, high K-values, age, low visual acuity, eye rubbing, atopic dermatitis (self-reported), and smoking. Odds ratios were calculated at each time point. Especially in the long-term, young age, high astigmatism (> 4.3 D), thin cornea (< 480 µm), poor initial visual acuity, and atopic dermatitis were risk factors for CXL failure in the sense of new progression after CXL. In contrast, smokers were more likely to belong to the responder group. No correlation between the outcome and initial level of K_max_ (Krumeich stage) was found. Additionally, there was no correlation with patient-reported eye rubbing. However, not every patient was asked about eye rubbing, making a final statement impossible.

**Fig. 3:**
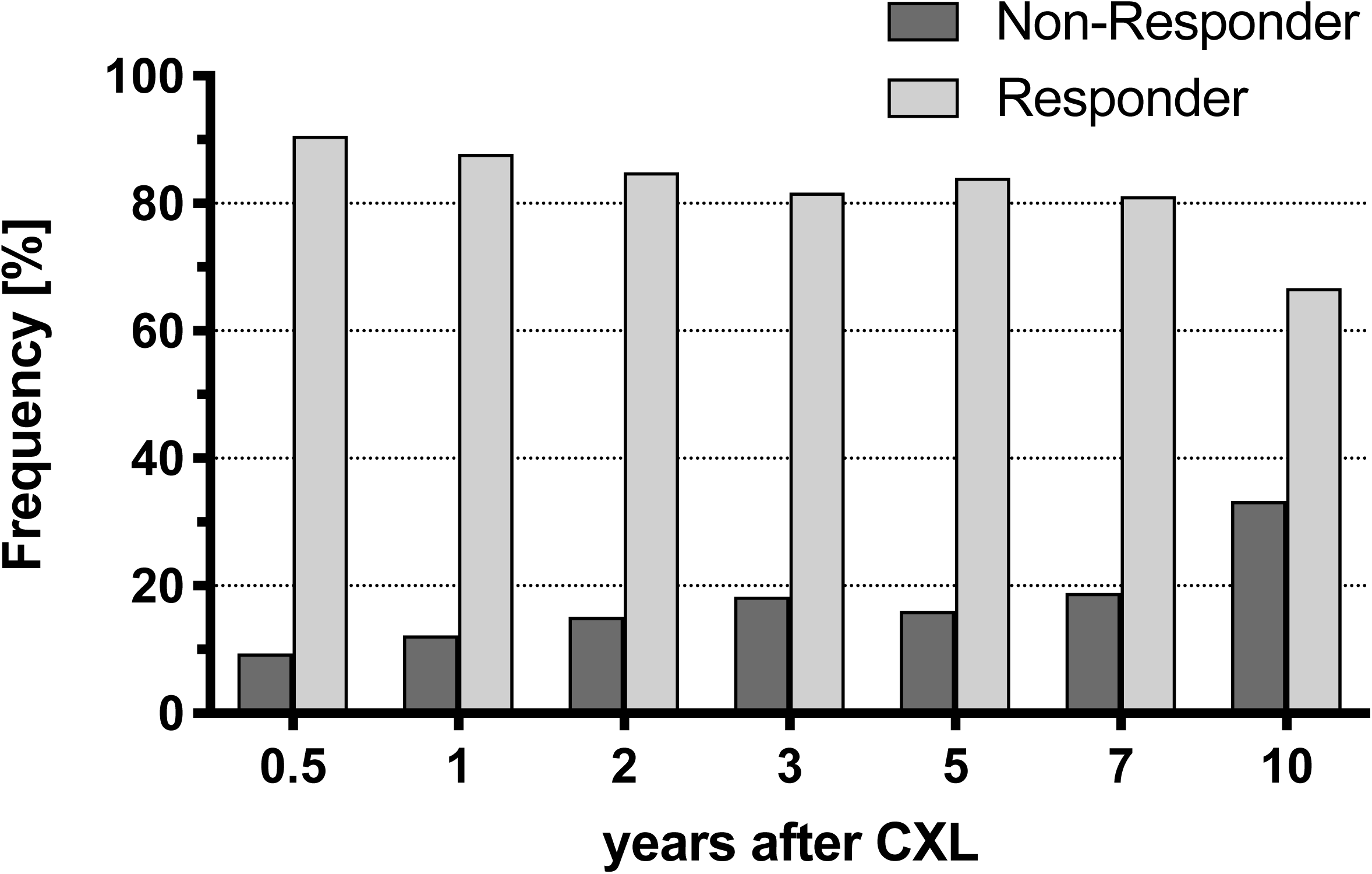
Rate of responders and non-responders over the entire follow-up time. Beyond 5 years postoperatively, a significant increase in the non-responder rate was observed

4 patients showed rapid keratoconus progression after CXL, and re-treatment was performed without complications after 3 years (one eye) and after 4 years (three eyes). In all re-treated patients, a stabilization of keratoconus was observed. All of them showed two risk factors for belonging to the non-responder group: two patients were young at the time of treatment (17 and 19 years of age), while the other two had high preoperative astigmatism >5 D.

No serious complications occurred after CXL. One year after treatment, mild haze could be observed in 14 % (14 of 103 eyes) of patients. After 10 years, 9 % (4 of 44 eyes) still showed haze. Corneal scars were present in 15 % (15 of 103 eyes) of the patients examined 1 year postoperatively, in 11 % (5 of 44 eyes) after 10 years. However, this did not lead to a loss of visual acuity in any case.

In the course of this study, 37 patients (28 %) underwent CXL also in the fellow eye, on average 13.5 ± 10.5 months after CXL of the first eye. 54 % of the fellow eyes were treated within the first year after CXL of the first eye, 87 % within 3 years. Because this study was designed to statistically compare patient-specific features, fellow eyes were not included in the analysis to avoid bias.

## Discussion

To date, there have been only three studies with a long-term follow-up of 10 years^12-14^, and one study with a mean follow-up of 11 years^15^. In line with these studies, our results confirm that CXL is an effective therapy for preventing keratoconus progression. Regarding changes in K_2_, we noted a stabilization of the disease from one year after treatment onwards, as shown by a K_2_ decrease of 0.85 D after 10 years. Theuring et al. and Raiskup et al. also observed a significant decrease in K_2_ in their 10-year results. At 4 D and 3.6 D, respectively, this was appreciably higher than in our study. Both studies also reported a significant decrease in K_max_ by 7 D and 6.2 D after 10 years^12,14^. Other studies with shorter follow-up periods have also reported a decrease in K_max_ after CXL treatment. O’Brart et al. reported a significant decrease of 0.4 D after 1 and 0.9 D after 5 and 7 years in a randomized double-blind study with 36 eyes of 36 patients^16^. Wittig-Silva et al. published the results of a prospective randomized trial with 100 eyes of 100 patients with progressive keratoconus in 2014. They also observed an improvement in K_max_ by 1 dpt after 3 years in the treatment group^17^. Our study showed a non-significant decrease in median K_max_ after 3 and 5 years, but then an increase in the postoperative course. K_max_, as measured by Pentacam in our study, represents the steepest curvature of the cornea at a single measurement point on the corneal surface. Therefore, it is highly susceptible to outliers and fluctuations. To assess changes in the corneal curvature after CXL, we advise to focus not only on K_max_ but also on the development of K_2_.

Regarding apical corneal thickness, we observed a tendency toward thickening beyond 5 years after surgery. Up to 5 years post-CXL, the median thickness was reduced and almost returned to baseline after 10 years. Caporossi et al. also reported an increase in corneal thickness during the 4-year postoperative period^11^.

Best-corrected visual acuity in our study stabilized in half and improved in one-third of the patients after three years. After 10 years, the mean CDVA improved significantly by one line (0.08 logMAR). This is in accordance with other studies reporting visual acuity improvement or stabilization 3 ^9,11,16,17^ or 10 years after treatment ^12,14,18^. However, beyond 5 years after treatment we observed a decrease in the proportion of patients with improved visual acuity, whereas the proportion of patients with vision loss increased slightly from 2 years onwards. Recently, Vinciguerra et al. reported long-term safety and efficacy of CXL for the treatment of keratoconus. The authors examined 27 eyes of 22 patients with a mean follow-up of 11 years, one eye had a follow-up time of 13 years. They also observed stabilization of visual acuity in their long-term results, as well as a significant decrease in the central keratoconus index and anterior surface curvature^15^.

To identify the factors that increase the likelihood of therapy failure, we divided the patients into responders and non-responders. To clearly discriminate between the groups, we chose a >2 D gap in postoperative ΔK_max_ between both groups. The proportion of non-responders increased beyond 5 years postoperatively. Thus, in the long-term, a new progression of the disease seems to become more probable.

Within the first 3 years after CXL, responders were younger than non-responders at the time of treatment. Caporossi et al. also reported that young patients (aged <19 years) benefited from the therapy^10^. However, from 5 years onwards, we observed a reversal: responders were older than non-responders. This can be explained by the fact that patients who were already older at the time of surgery had a higher chance of a natural therapy-independent disease stabilization. Younger patients, on the other hand, seem to particularly benefit from early therapy; however, in the long term, they show an increased risk of new progression. Regarding preoperatively recorded astigmatism, we observed an increased risk of CXL failure with an initially high astigmatism. This was also reflected in the results of Tuft et al., in which high astigmatism was associated with an increased risk of postoperative progression^19^.

Preoperative apical corneal thickness also influences the CXL outcomes. Thus, it was always higher in responders (average thickness >480 µm) than in non-responders.

The mean preoperative CDVA was better in responders than in non-responders. A better initial visual acuity of mean 0.2 logMAR or 20/32 was associated with a higher chance of therapy success compared to a mean 0.3 logMAR or 20/40 in the non-responder group. In contrast, Koller et al. identified a good initial CDVA of more than 20/25 Snellen lines as a significant risk factor for CXL complications, i.e., losing 2 or more Snellen lines^20^. The 20/25 value in this study, however, is considerably higher than the CDVA of our patients, and therefore does not contradict our results.

As expected and also stated by Lenk et al.^21^, atopic dermatitis was associated with a higher risk of therapy failure in our study. As previously described by Spoerl et al.^22^, a positive effect of smoking was also observed in our patients.

Although young age, high astigmatism, lower visual acuity, and atopic dermatitis have been identified as risk factors for non-response to CXL, these patients should not be excluded from CXL therapy. On the contrary, from our data, especially from the low complication rate and good results with re-treatment, we conclude that CXL should also be offered to patients at risk for non-response. These patients should be monitored for a longer period, and the procedure should be repeated in cases of keratoconus progression after CXL.

We repeated CXL in 4 patients without any complications. All of these 4 patients showed preoperative risk factors for being non-responders. After re-treatment, the disease stabilized in all eyes.

A limitation of this study is the possible distortion of the results due to the decreasing number of patients. Biased patient selection, especially in the long term, must be considered. For example, it is possible that particularly satisfied or dissatisfied patients no longer attended the follow-up examinations. However, the consistent pairwise comparison between pre- and postoperative data pairs for each individual time point, as applied in this study, renders the statistical analysis robust against intermittent dropouts and complete loss to follow-up.

In conclusion, these results demonstrate the efficacy of CXL for progressive keratoconus over a long-term follow-up period. However, beyond 5 years after treatment, the risk of progression increases, especially in patients younger than 30 years of age. Therefore, we recommend examining patients at regular intervals, especially beyond 5 years post CXL, to recognize and re-treat a progression early.

## Data Availability

All data produced in the present study are available upon reasonable request to the authors.

## Acknowledgments

The authors are grateful for the invaluable advice of the Institute for Clinical Epidemiology and Biometry (IKE-B) at the University of Würzburg in study planning and statistical analysis.

## Figure Legends

**Supplementary Fig. 1**. Development of parameters indicating keratoconus progression after corneal collagen cross-linking (CXL), for each time point postoperatively in comparison to the preoperative values. This allows plotting box blots with median, interquartile range, 25-75%-range and outliers.

(A) Median K_2_ change: Up to 1 year postoperatively, a slight increase of curvature was observed. Beyond 1 year, K_2_ decreased by up to 0.85 D at 10 years (*: p < 0.05, all other data points: not significant).

(B) Median K_max_ change: Fluctuations without significant changes were observed over the entire follow-up period.

(C) Median corneal thickness at the apex: Beyond 1 year postoperatively, a significant stabilization without any further thinning tendency was observed (***: p<0,001, **p < 0.005, *: p < 0.05).

(d) Change of median corrected distance visual acuity (CDVA): Beyond 2 years after treatment, an improvement of CDVA by about one line was observed (***: p<0,001, **p < 0.005, *: p < 0.05, n.s., not significant).

