## Supplementary figures and images for "Long-term outcome of corneal collagen crosslinking with riboflavin and UV-A irradiation for keratoconus"

### Supplementary Figure 1

#### Supplementary Figure 1

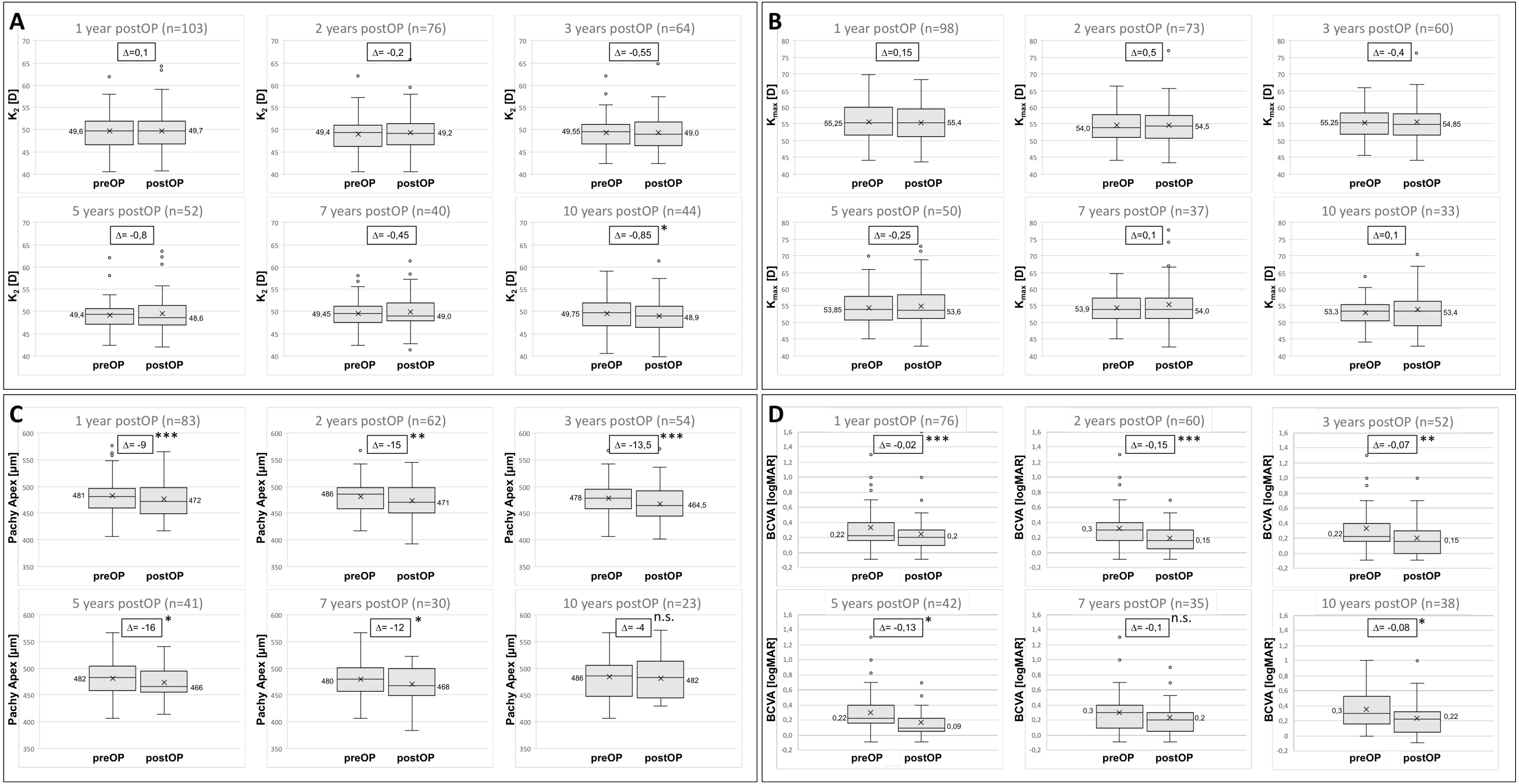
